# A study of risk analysis and prognosis models for the mortality of sepsis based on real-world data in China

**DOI:** 10.1101/2020.07.19.20151738

**Authors:** Yu Lu, Qing Kong, Jing Li, Tao Jiang, Zihui Tang

## Abstract

**Background:** The study aimed to explore the factors associated with the mortality of sepsis and to develop prognosis models for predicting outcomes based on real-world data in China. Methods: Data regarding sepsis patients’ medical records were extracted from the hospital information systems in four hospitals. The data included general information, laboratory tests, score systems, and supportive treatment for sepsis. In total, 507 medical records with complete data were available for data analysis. Multiple variable regression (MR) analysis used to explore associations, and to develop prognosis models

**Results:** The mortality of sepsis was 0.3124 in the total sample. A univariate analysis indicated 23 variables significantly associated with the mortality of sepsis (p <0.05 for all). The MLR analysis showed independent and significant variables of age, GCS, SOFA, shock, breath rate, TBIL, CHE, BUN, LAC, OI, HCO3-, IMV, and ALB (P <0.05 for all). Prognosis models have a high predictive performance (AUC = 0.885, 95% CI: 0.854–0.917 in model2). Conclusion: The study showed evidence of independent and significant factors associated with the mortality of sepsis, including age, GCS, SOFA, septic shock, breath rate, TBIL, CHE, BUN, LAC, OI, HCO_3_^-^, IMV, and ALB. Prognosis models with a high performance were developed. (Trial registration: ClinicalTrials.gov Identifier: NCTNCT03883061 retrospectively registered 19 Mar 2019.)

## Background

Severe sepsis is a potentially life-threatening complication of an infection, which could lead to chemicals being released into the bloodstream to fight the infection and the triggering of inflammatory responses throughout the body. This inflammation can trigger a cascade of changes that can damage multiple organ systems, causing them to fail [1].Severe sepsis is a leading cause of death in China and the most common cause of death among critically ill patients in intensive care units (ICUs) [2, 3]. Generally, the hospital mortality of sepsis is more than 30%. Most of the risk factors of sepsis are related to infection risk. For example, age, gender, and the increased burden of chronic health conditions are critical risk factors for the disease [3]. More importantly, most of the risk factors concerning vital signs contributed to the mortality of sepsis, such as blood pressure, shock status, or necessary vital signs to support treatment [3]. Unfortunately, it was inconsistent with the results of the risk related to the mortality of sepsis [1-6]. It is crucial for physicians to understand the risks for the mortality of sepsis to predict the outcome of sepsis in hospital.

Risk models or prognosis models that combine significant risk factors with mathematical models to estimate the mortality of sepsis are needed to aid in risk stratification and to identify high-risk individuals who may benefit from intensive interventions. Our previous studies created several predictive or risk models to predict the prevalence of diabetic complications [7, 8], the benefits of which include the prevention of diabetic complications. Predicting the mortality of sepsis should be more important for physicians to treat patients in hospital. However, few reports have assessed risk or prognosis models for the mortality of sepsis in China. Evidence from a real-world study is considered more representative of subject characteristics, making clinical practice more effective than randomized clinical trials (RCTs) [9, 10]. Real-world evidence is derived from real-world data (RWD) concerning the outcomes of heterogeneous patients in real-world practice settings [10]. Fortunately, information technology methods could effectively manage heterogeneous, multiple-source, and massive-scale RWD [11].

However, to the best of our knowledge, in Chinese sepsis patients, there have been no reports of risk factors and clinical risk models or prognosis models to predict the mortality of sepsis based on RWD. Clinical risk or prognosis models with high sensitivity and specificity are needed to predict the mortality of sepsis and to alert clinicians to the need for further treatment or clinical prediction. The purpose of the study was to explore the risk factors associated with the prognosis of sepsis and to create prognosis or risk models for predicting the mortality of sepsis based on RWD.

## Methods

### Study design and sample

Our group conducted a real-world study to evaluate the associated risk factors and create risk models for the mortality of sepsis in China. This was a multi-center, retrospective, observational study based on electronic medical information, including a hospital information system (HIS), electronic medical records (EMRs), and laboratory information system (LIS). In this study, between 2016and 2018, all EMRs on sepsis were collected from the emergency department ICUs in four hospitals - the hospital affiliated with South China University, the hospital affiliated with Jining Medical University, Hunan People’s Hospital, and Huaihua People’s Hospital. The inclusion criteria were as follows: diagnoses satisfying the diagnostic criteria of sepsis with detailed medical records, a first final diagnosis of sepsis, and subjects being over the age of 16. The exclusion criteria were as follows: unclear diagnoses or diagnoses not satisfying the diagnostic criteria of sepsis, unclear or incomplete medical records, and sepsis patients not satisfying the age criteria.

### Data collection and preparation

RWD regarding sepsis patients’ medical records were extracted from the HIS and EMRs of four hospitals. The data consist of general information, a laboratory test, a score system, and supportive treatment for sepsis patients.

The general information consists of age, heart rate, breathe rate, systolic blood pressure (SBP), and diastolic blood pressure (DBP). The parameters of the laboratory test include a regular blood test: white blood cell (WBC), hemoglobin (HB), and blood platelet (PLT); live function test: albumin (ALB), total bilirubin (TBIL), alanine aminotransferase (ALT), and choline esterase (CHE); renal function test: blood urea nitrogen (BUN) and serum creatinine (SCR); the parameters of a blood gas analysis: lactic acid (LAC), potential of hydrogen (PH), partial pressure of CO_2_ (PaCO_2_), partial pressure of O_2_ (PaO_2_), oxygenation index (OI), and bicarbonate (HCO_3_^-^); inflammation assays: C-reactive protein (CRP) andprocalcitonin (PCT); and plasma blood glucose (PBG).The parameters of the score system include the Glasgow Coma Scale (GCS)and sequential organ failure assessment (SOFA). The medical history records included diabetes mellitus (DM), chronic obstructive pulmonary disease (COPD), hypertension (HTN), and surgery history (SUR). The supportive treatment for sepsis patients concerned fluid infusion in 3 hours (FI3), fluid infusion in 6 hours (FI6), invasive mechanical ventilation (IMV), sedative medication use (SED), hypertensive agent use (HAU), glucocorticoid use (GLU),albumin transfusion (ALBT),plasma transfusion (PLAT),and the first time to use antibiotics (ATBT).

Data were transferred to structure records with 50 attributes, which were loaded to a data analysis platform with Python 3.6. The data analysis preparations were performed for data cleaning, deleting duplication records, deleting outliers, reconstructing missing data, etc. In total, 507 medical records on sepsis with complete data were available for future data analysis.

### Measurement and definition

BP values were the means of two physician-obtained measurements taken from the left arm of the seated participant. Laboratory tests concerning live function and renal function were measured using an enzymatic method with a chemical analyzer (ROHE COBAS 702). The regular blood test was conducted using a Sysmex XN2000analyzer, and the blood gas analysis was conducted using a Medica Easy BloodGasanalyzer. The unitized laboratory test protocol and assays were used according to the measurements in a multi-centre medical laboratory test platform. The day-to-day and inter-assay coefficients of variation at the central laboratory in our hospital for all analyses were between 1% and 3%.

The GCS score was calculated based on three aspects of responsiveness: eye-opening, motor, and verbal responses based on guidelines[12]. The SOFA score was computed based on the guidelines of Sequential Organ Failure Assessment (SOFA) score [13]. The conditions of sepsis patients require supportive treatment, including IMV, SED, HAU, GLU, PLAT, ALBT, and ATBT, which are performed according to the strategy of classics guidelines of sepsis[14].

Septic shock was defined as a subset of sepsis in which underlying circulatory and cellular metabolism abnormalities are profound enough to increase mortality substantially [15, 16].DM was defined by an oral glucose tolerance test (OGTT) or the use of insulin or hypoglycemic medications. COPD was defined as a common, preventable, and treatable disease characterized by persistent respiratory symptoms and airflow limitation based on guidelines [17]. HTN was defined as a BP of ≥140/90mmHgor a history of hypertension medication. The definition of sepsis was organ dysfunction caused by a dysregulated host response to infection according to the Third International Consensus Definitions for Sepsis and Septic Shock (Sepsis-3 definition)[15, 18]. The study outcome concerning the mortality of sepsis was defined as death within the 28-day period or survival at 28 days after ICU admission as endpoints.

### Data analysis

In this study, the main procedure of data analysis was as follow: 1) descriptive statistics for subjects, 2) a univariate analysis to explore the factors associated with the outcome, 3) an MR analysis to include candidate risk factors for the outcome, and 4) prognosis model development and a performance analysis of the mortality of sepsis.

Continuous variables were described as the mean ± standard deviation or median, unless stated otherwise. Meanwhile, categorical variables were described as proportionate for a particular category. Differences in variables among sepsis patients grouped by gender were determined by a one-way analysis of variance. Among the groups, differences in properties were detected using the χ^2^test. Tests were two-sided, and a p-value of < 0.05 was considered significant. Univariate logistic regression (ULR) analyses were performed to include 50 attributes for exploring candidate factors associated with the mortality of sepsis. In addition, to enhance the power to explore associated factors, continuous candidate variables were categorized as categorical variables using a ULR model to detect associations (please see supplementary file 1). The categorized algorithms were based on the reference value of continuous variables and relationship of mortality of sepsis. A multiple variable logistic regression (MLR) analysis was conducted to control potential confounders to determine independent and significant variables associated with the outcome. Moreover, MLR analyses with backward stepwise methods were performed to find the best-fit model to include independent and significant risk factors associated with the outcome.

Prognosis models for predicting the mortality of sepsis were developed. Briefly, the protocol of prognosis model development and performance analysis was as follows: 1) development of a prognosis formula based on the final MLR model with categorical variables, 2) calculation of scores for sepsis patients with a prognosis formula, and 3) performance analysis for prognosis models. The MLR was used to calculate β-coefficients for known factors associated with the outcome. Variables significant at 0.1 were included in the MLR using stepwise backward elimination, with the mortality of sepsis as the dependent variable. A prognosis model was based on a sum score derived from the addition of the score for each variable to the risk model for each subject. One prognosis model including variables with categorical variables without GCS or SOFA (model 1) was developed, model 2 was developed to include independent associated factors with categorical variables with GCS without SOFA, and model 3 was developed to include SOFA without GCS. A receiver-operating characteristic (ROC) curve and area under the curve (AUC) were produced. The sensitivity and specificity were calculated for each cutoff score. The maximum sum of the sensitivity and specificity (Youden index) at a cutoff score was taken as the optimum. The optimum cutoff score in the three models was evaluated to calculate the positive predictive value (PPV), negative predictive value (NPV), positive likelihood ratio (PLR), and negative likelihood ratio (NLR). The parameters of Hosmer–Lemeshow (HL) statistics and accuracy were calculated for a model performance evaluation. Moreover, the cutoff scores for the low- and high-risk groups for the outcome were additionally evaluated to calculate the outcome and proportion of the total sample in the corresponding group. Results were analyzed using the Statistical Package for Social Sciences for Windows, version 24.0 (SPSS, Chicago, IL, USA).

## Results

### The descriptive statistics of subjects

The baseline characteristics of the 507 sepsis patients are shown in Table 1. In the total sample, the mean age was 64.43 years, and the proportion of females (n=240) was 47.33%. The average SBP and DBP was 115.14 mmHg and 69.04 mmHg, respectively. The mean ALB, TBIL, and LAC were 30.2 g/L, 27.49umol/L, and 32.66 mmol/L, respectively. In the total sample, the prevalence of shock, DM, COPD, and HTN was 0.4311, 0.2264, 0.1591, and 0.3281, respectively. The rate of IMV use and SED use was 0.3379 and 0.3399, respectively. The mortality of sepsis was 0.3124 in the total sample.

**Table 1:** Descriptive statistics of subjects

| Variable | Female (n=240) | Male (n=269) | Total (n=509) | P value |
| --- | --- | --- | --- | --- |
| General information |  |  |  |  |
| Age | 64.53±13.89 | 64.34±15.23 | 64.43±14.6 | 0.883 |
| Hear rate (bpm) | 104.83±22.23 | 108.5±24.99 | 106.77±23.78 | 0.082 |
| Breath rate (bpm) | 24.97±7.89 | 25.66±8.31 | 25.33±8.11 | 0.335 |
| SBP (mmHg) | 114.34±31.57 | 115.85±30.32 | 115.14±30.9 | 0.584 |
| DBP (mmHg) | 67.68±16.45 | 70.32±18.79 | 69.07±17.75 | 0.094 |
| Laboratory test |  |  |  |  |
| WBC ($\times 10^9/L$) | 14.66±10.2 | 14.43±9.94 | 14.54±10.05 | 0.798 |
| HB (g/L) | 112.83±70.72 | 120.22±53.86 | 116.73±62.43 | 0.183 |
| PLT ($\times 10^9/L$) | 135.27±106.58 | 145.86±108.39 | 140.87±107.56 | 0.268 |
| ALB (g/L) | 29.97±5.53 | 30.4±5.91 | 30.2±5.73 | 0.395 |
| TBIL (umol/L) | 26.05±29.56 | 28.78±31.27 | 27.49±30.48 | 0.313 |
| ALT (U/L) | 100.65±237.38 | 96.21±271.66 | 98.31±255.83 | 0.845 |
| CHE (U/L) | 4185.11±1843.12 | 3731.95±2018.55 | 3945.07±1948.85 | 0.019 |
| BUN (mmol/L) | 13.37±9.83 | 14.03±11.52 | 13.72±10.75 | 0.489 |
| SCR(umol/L) | 167.9±146.41 | 173.69±147.35 | 170.96±146.8 | 0.657 |
| LAC (mmol/L) | 44.95±502.25 | 21.71±178.04 | 32.66±368.15 | 0.478 |
| PH | 7.31±0.24 | 7.3±0.23 | 7.3±0.23 | 0.609 |
| PaCO<sub>2</sub> (mmHg) | 31.71±11.03 | 34.07±12.87 | 32.96±12.08 | 0.028 |
| PaO<sub>2</sub> (mmHg) | 85.33±19.98 | 79.71±21.03 | 82.35±20.71 | 0.002 |
| OI | 315.77±169.03 | 275.8±153.84 | 294.61±162.24 | 0.005 |
| HCO<sub>3</sub><sup>-</sup> (mmol/L) | 19.23±7.04 | 19.87±6.8 | 19.57±6.91 | 0.298 |
| CRP (mg/L) | 132.08±79.34 | 124.61±82.53 | 128.13±81.05 | 0.300 |
| PCT (ug/L) | 41.54±57.98 | 31.88±37.04 | 36.41±48.19 | 0.024 |
| PBG (mmol/L) | 9.72±5.92 | 8.62±4.11 | 9.13±5.06 | 0.015 |
| Score system |  |  |  |  |
| GCS | 13.38±2.95 | 13.45±2.91 | 13.41±2.93 | 0.795 |
| SOFA | 8.76±6.63 | 9.39±7.33 | 9.09±7.01 | 0.313 |
| Medical history |  |  |  |  |
| Shock (yes) | 0.4625 | 0.403 | 0.4311 | 0.176 |
| DM (yes) | 0.2762 | 0.1822 | 0.2264 | 0.012 |
| COPD (yes) | 0.0958 | 0.2156 | 0.1591 | 0.000 |
| HTN (yes) | 0.3583 | 0.3011 | 0.3281 | 0.170 |
| Surgery (yes) | 0.0333 | 0.0855 | 0.0609 | 0.014 |
| Support treatment |  |  |  |  |
| FI3 (ml) | 1178.83±722.54 | 1125.28±723.6 | 1150.53±722.88 | 0.405 |
| FI6 (ml) | 1715.33±894.01 | 1710.32±1093.02 | 1712.68±1003.15 | 0.955 |
| IMV (yes) | 0.3167 | 0.3569 | 0.3379 | 0.338 |
| SED (yes) | 0.3458 | 0.3346 | 0.3399 | 0.789 |
| HAU (yes) | 0.5105 | 0.4796 | 0.4941 | 0.487 |
| GLU (yes) | 0.2583 | 0.2788 | 0.2692 | 0.603 |
| PLAT (yes) | 0.1958 | 0.1487 | 0.1709 | 0.158 |
| ALBT (yes) | 0.5833 | 0.4758 | 0.5265 | 0.015 |
| ATBT (hours) | 1.95±2.15 | 2.24±2.28 | 2.1±2.22 | 0.138 |
| Outcome |  |  |  |  |
| Mortality | 0.300 | 0.3234 | 0.3124 | 0.569 |

Note: SBP - systolic pressure, DBP - diastolic pressure, WBC - white blood cell , HB - hemoglobin, PLT - blood platelet, ALB - albumin, TBIL - total bilirubin, ALT - alanine aminotransferase, CHE - choline esterase, BUN - blood urea nitrogen, SCR - serum creatinine, LAC - lactic acid, PH - potential of hydrogen, PaCO<sub>2</sub> - partial pressure of CO<sub>2</sub>, PaO<sub>2</sub> - partial pressure of O<sub>2</sub>, OI - oxygenation index, HCO<sub>3</sub><sup>-</sup> - bicarbonate, CRP - C-reactive protein, PCT - procalcitonin, PBG - plasma blood glucose, GCS - Glasgow Coma Scale, SOFA - sequential organ failure assessment, DM - diabetes mellitus, COPD - chronic obstructive pulmonary disease, HTN - hypertension, SUR - surgery history, FI3 - fluid infusion in 3 hours, FI6 - fluid infusion in 6 hours, IMV - invasive mechanical ventilation, SED – sedative medication use, HAU - Hypertensive agent use -, GLU - glucocorticoid, ALBT - albumin transfusion, PLAT - plasma transfusion, ATBT - the first time to use antibiotics

### Univariate analyses of the mortality of sepsis

The ULR analysis, which included general information parameters, showed that heart rate and breath rate were significantly associated with the outcome (p = 0.005and p=0.043 for breath rate, Table 2), respectively. The parameters of PLT, ALB, CHE, BUN, SCR, LAC, PH, OI, and HCO_3_^-^were significantly associated with the mortality of sepsis (P <0.05 for all, Table 2), respectively. The GCS was significantly associated with the mortality of sepsis (P <0.0001, Figure 1), and there was a negative correlation with the outcome. Sepsis patients with shock were significantly greater than patients without shock (0.19 vs. 0.475, p<0.0001). Concerning the parameters of supportive treatment, IMV and SED use were significantly associated with the outcome (p <0.0001 for IMV and p<0.001 for SED, Table 2), respectively. In addition, after categorizing continuous variables to categorical variables (Figure 1 and Figure 2), additional parameters, including SBP, TBIL, ALT, PaCO_2_, PaO_2_, PCT, PBG, SOFA, and FI6,were significantly associated with the outcome (p<0.05 for all, Table 3), respectively.

**Table 2:** Univariate Logistic analysis for candidate associated factor associated with morality of sepsis

| Variable | $\beta$ | S.E | *P* value | OR | 95% CI |
| --- | --- | --- | --- | --- | --- |
| General information |  |  |  |  |  |
| Age | 0.027 | 0.012 | 0.025 | 1.027 | 1.003-1.052 |
| Gender | 0.387 | 0.305 | 0.205 | 1.472 | 0.810-2.678 |
| Heart rate | 0.018 | 0.006 | 0.005 | 1.018 | 1.005-1.031 |
| Breath rate | 0.031 | 0.015 | 0.043 | 1.031 | 1.001-1.062 |
| SBP | 0.003 | 0.005 | 0.513 | 1.003 | 0.993-1.013 |
| DBP | 0.004 | 0.008 | 0.577 | 1.005 | 0.989-1.021 |
| Laboratory test |  |  |  |  |  |
| WBC | 0.001 | 0.015 | 0.944 | 1.001 | 0.972-1.031 |
| HB | 0.033 | 0.059 | 0.575 | 1.034 | 0.921-1.16 |
| PLT | 0.003 | 0.002 | 0.049 | 1.003 | 1.000-1.006 |
| Alb | -0.061 | 0.028 | 0.029 | 0.941 | 0.891-0.994 |
| TBIL | 0.003 | 0.006 | 0.562 | 1.003 | 0.992-1.015 |
| ALT | 0.001 | 0.001 | 0.346 | 1.001 | 0.999-1.002 |
| CHE | 0.001 | 0.001 | 0.001 | 1.001 | 1.000-1.001 |
| BUN | 0.065 | 0.019 | 0.001 | 1.067 | 1.028-1.108 |
| Scr | 0.002 | 0.001 | 0.023 | 1.002 | 1.000-1.005 |
| LAC | 0.222 | 0.053 | <0.0001 | 1.248 | 1.126-1.384 |
| PH | -3.892 | 1.584 | 0.014 | 0.020 | 0.001-0.455 |
| PaCO<sub>2</sub> | -0.026 | 0.017 | 0.115 | 0.974 | 0.942-1.007 |
| PaO<sub>2</sub> | -0.003 | 0.004 | 0.47 | 0.997 | 0.990-1.005 |
| OI | -0.003 | 0.001 | 0.009 | 0.997 | 0.994-0.999 |
| HCO<sub>3</sub><sup>-</sup> | -0.096 | 0.031 | 0.002 | 0.908 | 0.855-0.965 |
| CRP | -0.003 | 0.002 | 0.168 | 0.997 | 0.992-1.001 |
| PCT | -0.003 | 0.004 | 0.449 | 0.997 | 0.990-1.005 |
| PBG | -0.020 | 0.041 | 0.619 | 0.98 | 0.904-1.062 |
| Score system |  |  |  |  |  |
| GCS | -0.358 | 0.072 | <0.0001 | 0.699 | 0.607-0.804 |
| SOFA | 0.143 | 0.047 | 0.003 | 1.153 | 1.051-1.265 |
| Medical history |  |  |  |  |  |
| Shock | 1.351 | 0.220 | <0.0001 | 3.853 | 2.591 - 5.724 |
| DM | -0.651 | 0.418 | 0.119 | 0.522 | 0.230-1.183 |
| COPD | 0.642 | 0.363 | 0.077 | 1.900 | 0.933-3.867 |
| HTN | 0.135 | 0.340 | 0.691 | 1.145 | 0.588-2.231 |
| SUR | 0.739 | 0.486 | 0.128 | 2.095 | 0.808-5.43 |
| Support treatment |  |  |  |  |  |
| FI3 | 0.001 | 0.001 | 0.068 | 1.000 | 0.999-1.000 |
| FI6 | 0.001 | 0.001 | 0.299 | 1.000 | 0.999-1.000 |
| IMV | 1.842 | 0.364 | <0.0001 | 6.311 | 3.092-12.878 |
| SED | 1.115 | 0.331 | 0.001 | 3.049 | 1.594-5.830 |
| HAU | -0.093 | 0.298 | 0.756 | 0.911 | 0.509-1.634 |
| GLU | 0.273 | 0.377 | 0.469 | 1.314 | 0.628-2.749 |
| ALBT | 0.562 | 0.309 | 0.069 | 1.754 | 0.958-3.211 |
| PLAT | 0.469 | 0.425 | 0.270 | 1.598 | 0.694-3.677 |
| ATBT | -0.022 | 0.056 | 0.690 | 0.978 | 0.876-1.091 |

Note: SBP - systolic pressure, DBP - diastolic pressure, WBC - white blood cell, HB - hemoglobin, PLT - blood platelet, Alb - albumin, TBIL - total bilirubin, ALT - alanine aminotransferase, CHE - choline esterase, BUN - blood urea nitrogen, Scr - serum creatinine, LAC - lactic acid, PH - potential of hydrogen, PaCO<sub>2</sub> - partial pressure of CO<sub>2</sub>, PaO<sub>2</sub> - partial pressure of O<sub>2</sub>, OI - oxygenation index, HCO<sub>3</sub><sup>-</sup> - bicarbonate, CRP - C-reactive protein, PCT - procalcitonin, PBG - plasma blood glucose, GCS - Glasgow Coma Scale, SOFA - sequential organ failure assessment, DM - diabetes mellitus, COPD - chronic obstructive pulmonary disease, HTN - hypertension, SUR - surgery history, FI3 - fluid infusion in 3 hours, FI6 - fluid infusion in 6 hours, IMV - invasive mechanical ventilation, SED – sedative medication use, HAU - Hypertensive agent use -, GLU - glucocorticoid, ALBT - albumin transfusion, PLAT - plasma transfusion, ATBT - the first time to use antibiotics

**Table 3:** Univarite analysis for candidate risk factors of mortality of sepsis with categorized variable

| Variable | $\beta$ | S.E | *P* value | OR | 95% CI |
| --- | --- | --- | --- | --- | --- |
| General information |  |  |  |  |  |
| Age | 0.514 | 0.157 | 0.001 | 1.672 | 1.229 - 2.275 |
| Heart rate | 0.528 | 0.199 | 0.008 | 1.695 | 1.148 - 2.502 |
| Breath rate | 0.622 | 0.151 | 0.000 | 1.862 | 1.386 - 2.503 |
| SBP | 0.573 | 0.226 | 0.011 | 1.774 | 1.14 - 2.761 |
| DBP | 0.313 | 0.202 | 0.122 | 1.367 | 0.919 - 2.032 |
| Laboratory test |  |  |  |  |  |
| WBC | 0.306 | 0.132 | 0.021 | 1.358 | 1.048 - 1.76 |
| Hb | 0.489 | 0.204 | 0.017 | 1.630 | 1.093 - 2.432 |
| Plt | -0.003 | 0.194 | 0.986 | 0.997 | 0.681 - 1.458 |
| Alb | 0.716 | 0.149 | 0.000 | 2.047 | 1.529 - 2.74 |
| TBIL | 0.643 | 0.204 | 0.002 | 1.903 | 1.276 - 2.839 |
| ALT | 0.360 | 0.119 | 0.003 | 1.433 | 1.134 - 1.811 |
| CHE | 0.859 | 0.124 | 0.000 | 2.361 | 1.852 - 3.010 |
| BUN | 0.596 | 0.114 | 0.000 | 1.815 | 1.453 - 2.268 |
| SCR | 0.894 | 0.197 | 0.000 | 2.446 | 1.664 - 3.595 |
| LAC | 1.091 | 0.142 | 0.000 | 2.976 | 2.253 - 3.931 |
| PH | 1.033 | 0.230 | 0.000 | 2.808 | 1.789 - 4.408 |
| PaCO<sub>2</sub> | 0.379 | 0.118 | 0.001 | 1.461 | 1.160 - 1.840 |
| PaO<sub>2</sub> | 0.292 | 0.192 | 0.129 | 1.339 | 0.919 - 1.951 |
| OI | 0.787 | 0.260 | 0.002 | 2.196 | 1.319 - 3.656 |
| HCO<sub>3</sub><sup>-</sup> | 0.967 | 0.205 | 0.000 | 2.631 | 1.76 - 3.932 |
| CRP | 0.096 | 0.256 | 0.707 | 1.101 | 0.667 - 1.819 |
| PCT | 0.286 | 0.124 | 0.021 | 1.331 | 1.045 - 1.696 |
| PBG | 0.592 | 0.256 | 0.021 | 1.807 | 1.094 - 2.987 |
| Score system |  |  |  |  |  |
| GCS | 0.945 | 0.118 | 0.000 | 2.572 | 2.042-3.239 |
| SOFA | 0.516 | 0.118 | 0.000 | 1.675 | 1.329 - 2.111 |
| Support treatment |  |  |  |  |  |
| FI3 | 0.028 | 0.138 | 0.838 | 1.029 | 0.785 - 1.348 |
| FI6 | 0.212 | 0.106 | 0.046 | 1.236 | 1.004 - 1.521 |
| ATBT | 0.361 | 0.216 | 0.095 | 1.434 | 0.94 - 2.189 |

Note: SBP - systolic pressure, DBP - diastolic pressure, WBC - white blood cell, HB - hemoglobin, PLT - blood platelet, Alb - albumin, TBIL - total bilirubin, ALT - alanine aminotransferase, CHE - choline esterase, BUN - blood urea nitrogen, Scr - serum creatinine, LAC - lactic acid, PH - potential of hydrogen, PaCO<sub>2</sub> - partial pressure of co<sub>2</sub>, PaO<sub>2</sub> - partial pressure of o<sub>2</sub>, OI - oxygenation index, HCO<sub>3</sub><sup>-</sup> - bicarbonate, CRP - C-reactive protein, PCT - procalcitonin, GCS - Glasgow Coma Scale, SOFA - sequential organ failure assessment, PBG - plasma blood glucose, FI3 - fluid infusion in 3 hours, FI6 - fluid infusion in 6 hours, ATBT - the first time to use antibiotics

**Figure 1:**
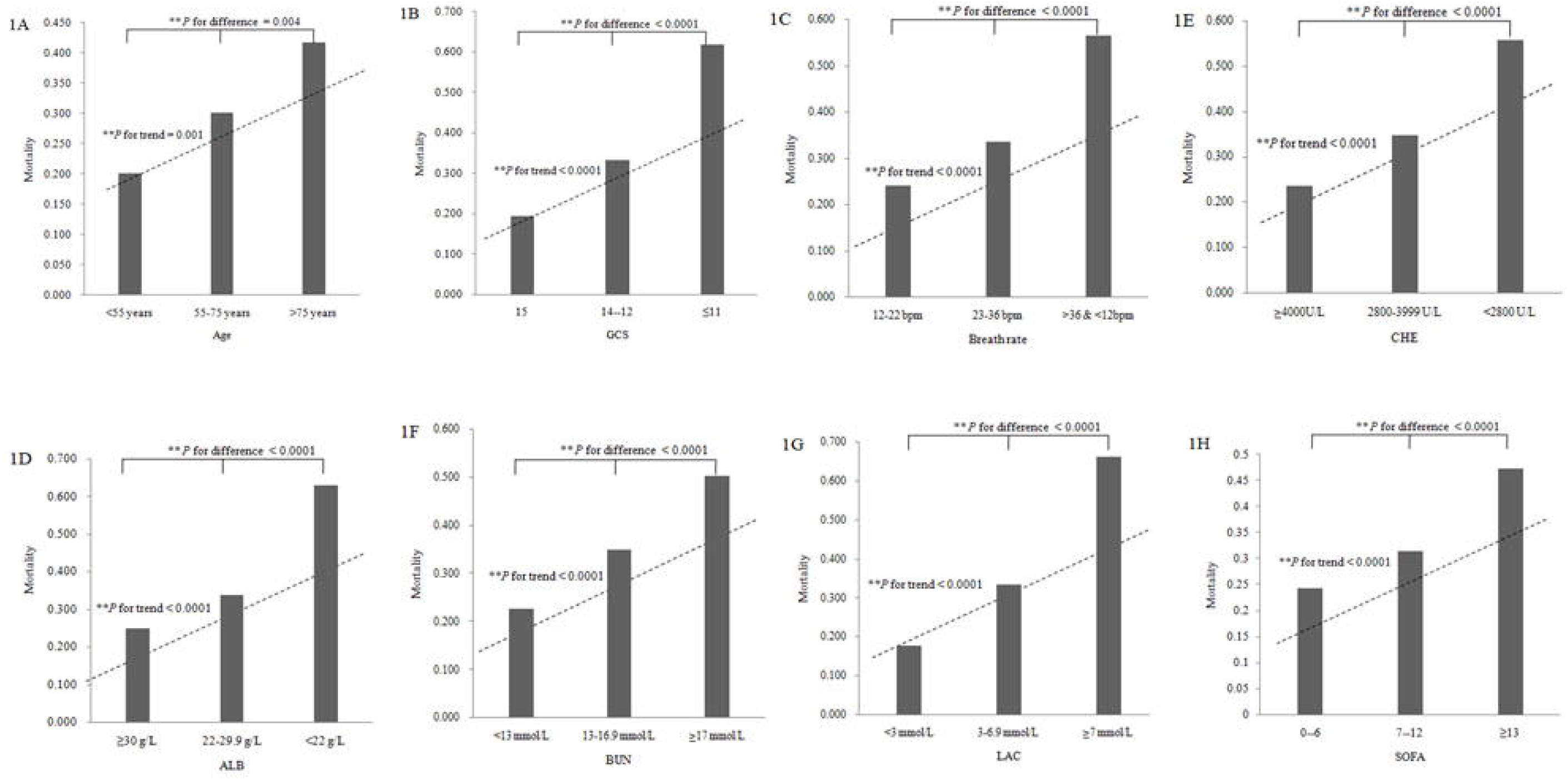
Comparison of mortality of sepsis among groups according to trinary variables. Figure 1A, the results of comparison of mortality of sepsis among groups according to Age (code 0: <55 years, code 1: 55-75 years, code 2: >75 years). The mortality of subjects with sepsis was 0.202, 0.302 and 0.417 in the three groups, respectively. There were significantly differences among the three groups (P for difference = 0.004 and P for trend = 0.001); Figure 1B, the results of comparison of mortality of sepsis among groups according to GCS (code 0: 15, code 1: 12-14, code 2: ≤11). The mortality of subjects with sepsis was 0.193, 0.333 and 0.618 in the three groups, respectively. There were significantly differences among the three groups (P for difference < 0.0001 and P for trend < 0.0001); Figure 1C, the results of comparison of mortality of sepsis among groups according to breath rate (code 0: 12-22 bpm, code 1: 23-36 bpm, code 2: >36 and <12 bpm). The mortality of subjects with sepsis was 0.241, 0.335 and 0.565 in the three groups, respectively. There were significantly differences among the three groups (P for difference < 0.0001 and P for trend < 0.0001); Figure 1D, the results of comparison of mortality of sepsis among groups according to ALB (code 0: ≥30g/L, code 1: 22-29g/L, code 2: <22g/L). The mortality of subjects with sepsis was 0.244, 0.333 and 0.651 in the three groups, respectively. There were significantly differences among the three groups (P for difference < 0.0001 and P for trend < 0.0001); Figure 1E, the results of comparison of mortality of sepsis among groups according to CHE (code 0: ≥4000U/L, code 1: 2800-3999U/L, code 2: <28000U/L). The mortality of subjects with sepsis was 0.235, 0.346 and 0.558 in the three groups, respectively. There were significantly differences among the three groups (P for difference < 0.0001 and P for trend < 0.0001); Figure 1F, the results of comparison of mortality of sepsis among groups according to BUN (code 0: <13 mmol/L, code 1: 13-16.9 mmol/L, code 2: ≥17 mmol/L). The mortality of subjects with sepsis was 0.232, 0.349 and 0.500 in the three groups, respectively. There were significantly differences among the three groups (P for difference < 0.0001 and P for trend < 0.0001); Figure 1G, the results of comparison of mortality of sepsis among groups according to LAC (code 0: <3 mmol/L, code 1: 3-6.9 mmol/L, code 2: ≥7 mmol/L). The mortality of subjects with sepsis was 0.177, 0.352 and 0.671 in the three groups, respectively. There were significantly differences among the three groups (P for difference < 0.0001 and P for trend < 0.0001); Figure 1H, the results of comparison of mortality of sepsis among groups according to SOFA (code 0: 0--6, code 1: 7--12, code 2: ≥13). The mortality of subjects with sepsis was 0.243, 0.315 and 0.473 in the three groups, respectively. There were significantly differences among the three groups (P for difference < 0.0001 and P for trend < 0.0001); GCS - Glasgow Coma Scale, ALB - albumin, CHE - choline esterase, BUN - blood urea nitrogen, LAC - lactic acid, SOFA - sequential organ failure assessment.

**Figure 2:**
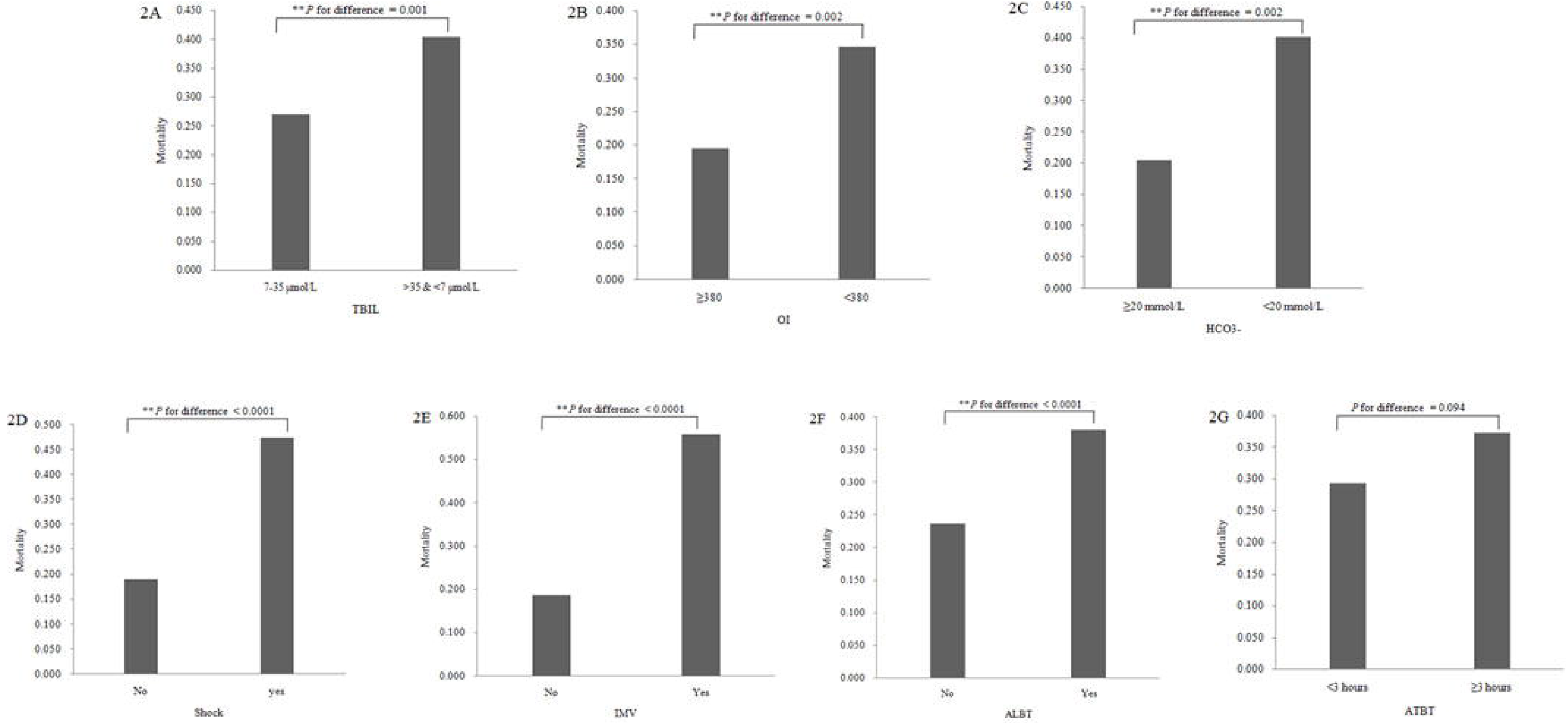
Comparison of mortality of sepsis between groups according to binary variables. Figure 2A, the results of comparison of mortality of sepsis between groups according to TBIL (code 0: 7-35 umol/L and code 1: ≥35 and <7 umol/L). The mortality of subject with sepsis was 0.270 and 0.413 in the two groups, respectively. There were significantly differences between the two groups (P for difference = 0.001); Figure 2B, the results of comparison of mortality of sepsis between groups according to OI (code 0: ≥ 380 and code 1: <380). The mortality of subject with sepsis was 0.195 and 0.347 in the two groups, respectively. There were significantly differences between the two groups (P for difference = 0.002); Figure 2C, the results of comparison of mortality of sepsis between groups according to HCO_3_^-^ (code 0: ≥ 20 mmol/L, code 1: <20 mmol/L). The mortality of subject with sepsis was 0.203 and 0.401 in the two groups, respectively. There were significantly differences between the two groups (P for difference = 0.002); Figure 2D, the results of comparison of mortality of sepsis among groups according to shock (code 0: no, code 1: yes). The mortality of subject with sepsis was 0.190 and 0.475 in the two groups, respectively. There were significantly differences between the two groups (P for difference < 0.0001); Figure 2E, the results of comparison of mortality of sepsis among groups according to IMV (code 0: no, code 1: yes). The mortality of subject with sepsis was 0.187 and 0.558 in the two groups, respectively. There were significantly differences between the two groups (P for difference < 0.0001); Figure 2F, the results of comparison of mortality of sepsis among groups according to ALBT (code 0: no, code 1: yes). The mortality of subject with sepsis was 0.237 and 0.381 in the two groups, respectively. There were significantly differences between the two groups (P for difference < 0.0001); Figure 2G, the results of comparison of mortality of sepsis among groups according to ATBT (code 0: <3 hours, code 1: ≥3 hours). The mortality of subject with sepsis was 0.293 and 0.373 in the two groups, respectively. There were not significantly differences between the two groups (P for difference = 0.094); TBIL - total bilirubin, OI - oxygenation index, IMV - invasive mechanical ventilation, ATBT - the first time to use antibiotics.

### Multiple variable analyses of mortality of sepsis

An MLR analysis with the backward stepwise method indicated the parameters of age, septic shock, breath rate, ALB, BUN, LAC, PH, and IMV in the final model 1 (Table 4). The parameters of age, GCS, shock, breath rate, CHE, BUN, LAC, PaCO_2_, and IMV were reported in the final model 2 (Table 4). After categorizing continuous variables as categorical variables (Figure 1 and Figure 2), in the categorical model, the variables of age, septic shock, breath rate, ALB, TBIL, CHE, BUN, LAC, OI, IMV, and ATBT were reported in the final model 1 (initial input variables without GCS, Table 5). In addition, the variables of age, GCS, septic shock, breath rate, TBIL, CHE, BUN, LAC, OI, HCO3-, IMV, ALBT, and ATBT were reported in the final model 2 (Table 5), and the variables of age, LAC, SOFA, and ATBT were reported in the final model 3 (Table 5).

**Table 4:** Multiple variable logistic analysis for risk factors associated with mortality of sepsis with continuous and categorical variables

| Model | Variable | $\beta$ | S.E | *P* value | OR | 95% CI |
| --- | --- | --- | --- | --- | --- | --- |
| Model 1 |  |  |  |  |  |  |
|  | Age | 0.027 | 0.010 | 0.006 | 1.028 | 1.008 - 1.048 |
|  | Shock | 0.904 | 0.276 | 0.001 | 2.471 | 1.439 - 4.243 |
|  | Breath rate | 0.053 | 0.016 | 0.001 | 1.055 | 1.021 - 1.089 |
|  | ALB | -0.065 | 0.025 | 0.009 | 0.937 | 0.892 - 0.984 |
|  | BUN | 0.050 | 0.016 | 0.002 | 1.051 | 1.019 - 1.085 |
|  | LAC | 0.142 | 0.046 | 0.002 | 1.153 | 1.052 - 1.262 |
|  | PH | 1.110 | 0.614 | 0.071 | 3.034 | 0.91 - 10.117 |
|  | IMV | 1.784 | 0.283 | 0.000 | 5.955 | 3.419 - 10.369 |
| Model 2 |  |  |  |  |  |  |
|  | Age | 0.025 | 0.010 | 0.015 | 1.025 | 1.005 - 1.046 |
|  | GCS | -0.209 | 0.053 | 0.000 | 0.811 | 0.731 - 0.900 |
|  | Shock | 0.675 | 0.287 | 0.019 | 1.964 | 1.119 - 3.450 |
|  | Breath rate | 0.044 | 0.017 | 0.009 | 1.045 | 1.011 - 1.081 |
|  | CHE | 0.000 | 0.000 | 0.006 | 1.000 | 1.000 - 1.001 |
|  | BUN | 0.039 | 0.016 | 0.015 | 1.040 | 1.008 - 1.073 |
|  | LAC | 0.146 | 0.047 | 0.002 | 1.157 | 1.055 - 1.270 |
|  | PaCO2 | -0.028 | 0.015 | 0.058 | 0.972 | 0.944 - 1.001 |
|  | IMV | 1.419 | 0.318 | 0.000 | 4.131 | 2.214 - 7.708 |
| Model 3 |  |  |  |  |  |  |
|  | Age | 0.025 | 0.008 | 0.002 | 1.026 | 1.009-1.042 |
|  | LAC | 0.197 | 0.036 | 0.000 | 1.217 | 1.134-1.306 |
|  | SOFA | 0.057 | 0.016 | 0.000 | 1.059 | 1.027-1.092 |
|  | ATBT | 0.101 | 0.048 | 0.037 | 1.106 | 1.006-1.216 |

Note: Initial input variables in model 1: Age, Shock, Heart rate, Breath rate, SBP, Alb, TBIL, ALT, CHE, BUN, Scr, LAC, PH, PaCO<sub>2</sub>, OI, HCO<sub>3</sub><sup>-</sup>, PCT, IMV, SED, GLU, ALBT, PLAT, ATBT; Initial input variables in model 2: Age, GCS, Shock, Heart rate, Breath rate, SBP, Alb, TBIL, ALT, CHE, BUN, SCR, LAC, PH, PaCO<sub>2</sub>, OI, HCO<sub>3</sub><sup>-</sup>, PCT, SOFA, IMV, SED, GLU, ALBT, PLAT, ATBT; Initial input variables in model 3: gender, age, LAC, PCT, PBG, SOFA, GLU, ALBT, ATBT. SBP - systolic pressure, DBP - diastolic pressure, WBC - white blood cell, HB - hemoglobin, PLT - blood platelet, ALB - albumin, TBIL - total bilirubin, ALT - alanine aminotransferase, CHE - choline esterase, BUN - blood urea nitrogen, Scr - serum creatinine, LAC - lactic acid, PH - potential of hydrogen, PaCO<sub>2</sub> - partial pressure of co2, PaO<sub>2</sub> - partial pressure of o2, OI - oxygenation index, HCO<sub>3</sub><sup>-</sup> - bicarbonate, CRP - C-reactive protein, PCT - procalcitonin, SOFA - sequential organ failure assessment, DM - diabetes mellitus, COPD - chronic obstructive pulmonary disease, HTN - hypertension, SUR - surgery history, PBG - plasma blood glucose, FI3 - fluid infusion in 3 hours, FI6 - fluid infusion in 6 hours, IMV - invasive mechanical ventilation, SED - sedative medication use, HAU - Hypertensive agent use -, GLU - glucocorticoid, ALBT - albumin transfusion, PLAT - plasma transfusion, ATBT - the first time to use antibiotics

**Table 5:**
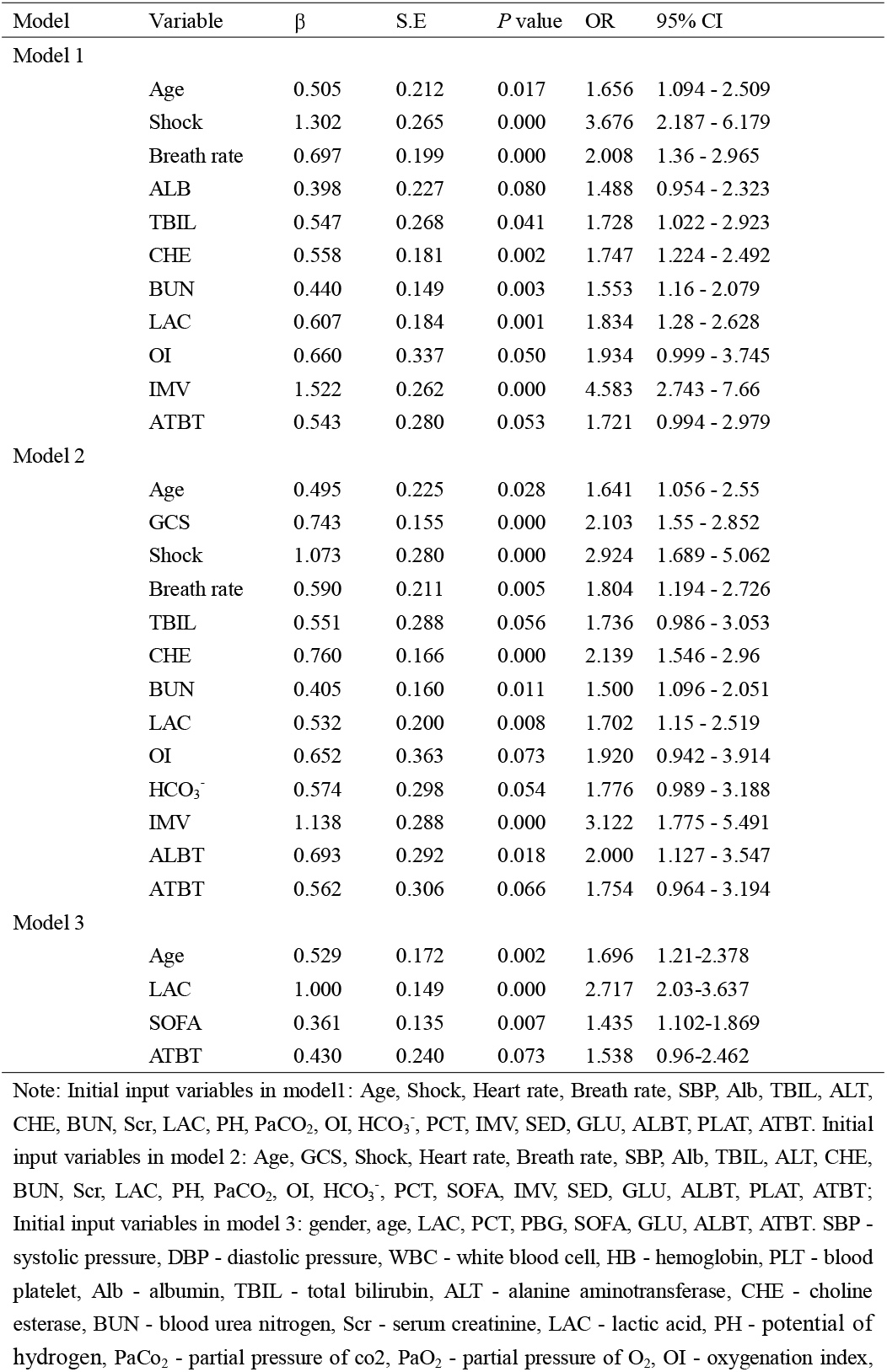
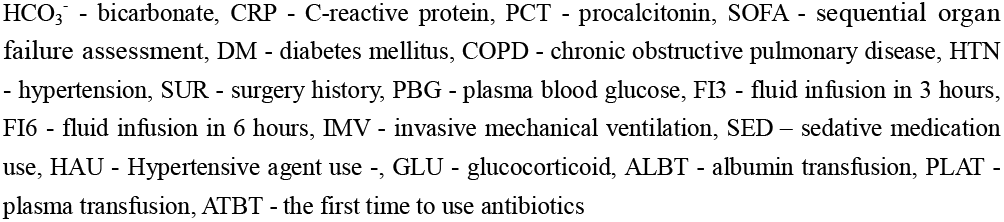
Multiple variable logistic analysis for risk factors associated with mortality of sepsis with categorized variables

### Prognosis models on mortality of sepsis

Three prognosis models were developed (please see Table 6). In model 1, the prognosis model with scores ranged between 0.00 and 10.33. The AUC was 0.865 (95% CI: 0.828–0.896, P < 0.001, Figure. 3), which was almost identical to that of the corresponding model. A cutoff score of 4.737 was optimum (sensitivity = 0.748; specificity = 0.814; Youden index = 0.563, Table 6). The PPV, NPV, PLR, and NLP were 0.644, 0.878, 4.034, and 0.309, respectively. The HL statistics and accuracy were 9.086 and 0.795, respectively. In model 2, the risk model with scores ranged between 0.00 and 10.33, and the AUC was 0.885 (95% CI: 0.854–0.917, P < 0.001, Figure. 3). A cutoff score of 5.427 was optimum (sensitivity = 0.801; specificity = 0.820; Youden index = 0.621, Table 6). The PPV, NPV, PLR, and NLP were 0.667, 0.902, 4.453, and 0.242, respectively. The HL statistics and accuracy were 4.977 and 0.814, respectively. In model 3, the risk model with scores ranged between 0.00 and 5.21, and the AUC was 0.736 (95% CI: 0.689 - 0.784, P < 0.001, Figure. 3).

**Table 6:** Predictive performance of risk model for mortality of sepsis

| Variable | Model 1 | Model 2 | Model 3 |
| --- | --- | --- | --- |
| AUC | 0.865 | 0.885 | 0.736 |
| 95% CI | 0.828-0.896 | 0.854-0.917 | 0.689-0.784 |
| Range | 0.0-10.33 | 0.0-11.63 | 0.0-5.21 |
| Cut point | 4.737 | 5.427 | 1.390 |
| Sensitivity | 0.748 | 0.801 | 0.780 |
| Specificity | 0.814 | 0.820 | 0.591 |
| Youden Index | 0.563 | 0.621 | 0.371 |
| PPV | 0.644 | 0.667 | 0.461 |
| NPV | 0.878 | 0.902 | 0.857 |
| PLR | 4.034 | 4.453 | 1.906 |
| NLR | 0.309 | 0.242 | 0.373 |
| HL Statistics | 9.086 | 4.977 | 7.637 |
| Accuracy | 0.795 | 0.814 | 0.649 |

Note: \*95% CI for AUC, AUC - Area under the receiver operating curve, PPV- positive predictive value, NPV - negative predictive value, PLR - positive likelihood ratio, NLR - negative likelihood ratio. HL statistics – Hosmer & Lemeshow statistics.
Formula of model 1: $0.505 \times \text{Age} + 1.302 \times \text{Shock} + 0.697 \times \text{Breath rate} + 0.398 \times \text{Alb} + 0.547 \times \text{TBIL} + 0.558 \times \text{CHE} + 0.440 \times \text{BUN} + 0.607 \times \text{LAC} + \text{OI} \times 0.660 + 1.522 \times \text{IMV} + 0.543 \times \text{ATBT}$ ;
Formula of model 2: $0.495 \times \text{Age} + 0.743 \times \text{GCS} + 1.073 \times \text{Shock} + 0.590 \times \text{Breath rate} + 0.551 \times \text{TBIL} + 0.760 \times \text{CHE} + 0.405 \times \text{BUN} + 0.532 \times \text{LAC} + 0.652 \times \text{OI} + 0.574 \times \text{HCO}_3^- + 1.138 \times \text{IMV} + 0.693 \times \text{ALBT} + 0.562 \times \text{ATBT}$ ;
Formula of model 3: $0.529 \times \text{Age} + 1.0 \times \text{LAC} + 0.361 \times \text{SOFA} + 0.43 \times \text{ATBT}$ .
SBP - systolic pressure, DBP - diastolic pressure, WBC - white blood cell, HB - hemoglobin, PLT - blood platelet, ALB - albumin, TBIL - total bilirubin, ALT - alanine aminotransferase, CHE - choline esterase, BUN - blood urea nitrogen, SCR - serum creatinine, LAC - lactic acid, PH - potential of hydrogen, PaCO<sub>2</sub> - partial pressure of co<sub>2</sub>, PaO<sub>2</sub> - partial pressure of o<sub>2</sub>, OI - oxygenation index, HCO<sub>3</sub><sup>-</sup> - bicarbonate, CRP - C-reactive protein, PCT - procalcitonin, SOFA - sequential organ failure assessment, DM - diabetes mellitus, COPD - chronic obstructive pulmonary disease, HTN - hypertension, SUR - surgery history, PBG - plasma blood glucose, FI3 - fluid infusion in 3 hours, FI6 - fluid infusion in 6 hours, IMV - invasive mechanical ventilation, SED – sedative medication use, HAU - Hypertensive agent use -, GLU - glucocorticoid, ALBT - albumin transfusion, PLAT - plasma transfusion, ATBT - the first time to use antibiotics, GCS - Glasgow Coma Scale.

**Figure 3:**
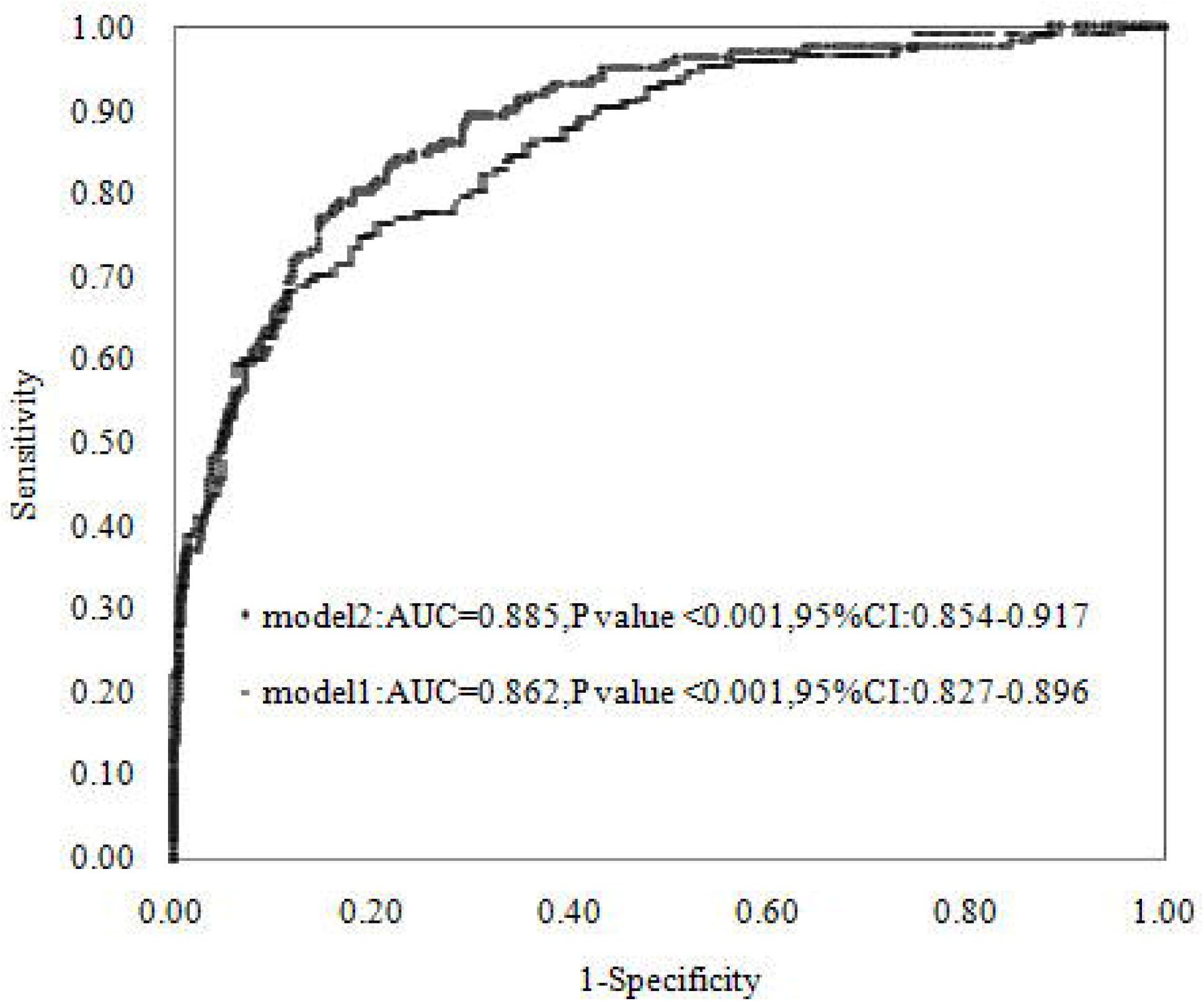
Receiver operating characteristic curves showed the performance of risk model 1, model 2 and model 3 in predicting mortality of sepsis. The 95% confidence interval (CI) for the AUC is shown. AUC - area under the curve.

Performance analyses of the low- and high-risk groups’ scores for the mortality of sepsis were conducted using the three models. In model 1, the cutoff score of 3.204 was set to the low-risk group (sensitivity = 0.950; specificity = 0.484; Youden index = 0.434, Table 7). The proportion of sepsis patients with a score less than the cutoff score of 3.204 was 0.347, having 0.045 of mortality. The cutoff score of 6.309 was set to the high-risk group. The proportion of sepsis patients with a score greater than the cutoff score of 6.309 was 0.135, having 0.912 of mortality. In model 2, the cutoff point for low risk was 4.398, and the proportion of subjects with a score less than the point was 0.455, having 0.049 of mortality; meanwhile, the cutoff point for high risk was 7.381, and the proportion of subjects with a score greater than the point was 0.127, having 0.903 of mortality. In model 3, the cutoff point for low risk was 0.841, and the proportion of subjects with score less than the point was 0.225, having 0.114 of mortality; meanwhile, the cutoff point for high risk was 2.925, and the proportion of subjects with score more than the point was 0.093, having 0.787 of mortality.

**Table 7:** Performance analysis of Low-risk and High-risk score group for sepsis

| Variable | Model 1 |  | Model 2 |  | Model 3 |  |
| --- | --- | --- | --- | --- | --- | --- |
|  | Low risk score | High risk score | Low risk score | High risk score | Low risk score | High risk score |
| Cut point | 3.204 | 6.309 | 4.398 | 7.381 | 0.841 | 2.925 |
| Sensitivity | 0.950 | 0.390 | 0.927 | 0.371 | 0.918 | 0.233 |
| Specificity | 0.484 | 0.983 | 0.625 | 0.982 | 0.291 | 0.971 |
| Youden Index | 0.434 | 0.373 | 0.553 | 0.353 | 0.209 | 0.204 |
| PPV | 0.453 | 0.910 | 0.526 | 0.904 | 0.291 | 0.784 |
| NPV | 0.955 | 0.782 | 0.950 | 0.777 | 0.908 | 0.738 |
| PLR | 1.841 | 22.421 | 2.475 | 20.954 | 0.915 | 8.075 |
| NLR | 0.104 | 0.621 | 0.116 | 0.640 | 0.225 | 0.790 |
| Accuracy | 0.628 | 0.794 | 0.719 | 0.793 | 0.485 | 0.742 |
| Mortality | 0.045 | 0.912 | 0.049 | 0.903 | 0.114 | 0.787 |
| Proportion | 0.347 | 0.135 | 0.455 | 0.127 | 0.225 | 0.093 |

Note: PPV- positive predictive value, NPV - negative predictive value, PLR - positive likelihood ratio, NLR - negative likelihood ratio. Model 1: the cutoff point for low risk was 3.204, and the proportion of subjects with score less than the point was 0.347, having 0.045 of mortality; and the cutoff point for high risk was 6.309, the proportion of subjects with score more than the point was 0.135, having 0.912 of mortality; Model2: the cutoff point for low risk was 4.398, the proportion of subjects with score less than the point was 0.455, having 0.049 of mortality; and the cutoff point for high risk was 7.381, the proportion of subjects with score more than the point was 0.127, having 0.903 of mortality; Model3: the cutoff point for low risk was 0.841, the proportion of subjects with score less than the point was 0.225, having 0.114 of mortality; and the cutoff point for high risk was 2.925, the proportion of subjects with score more than the point was 0.093, having 0.787 of mortality.

## Discussion

A real-world study was carried out to explore the associated factors and develop a prognosis model for the mortality of sepsis in China. RWE studies are increasingly becoming the normal clinical practice to ensure its significance [9, 10]. It is crucial to manage EMRs for this work and data analysis. The multicenter, retrospective, hospital-based observational study was designed to evaluate associations, providing the valuable experience of prognosis model development in a real-world study for clinical research. The understanding of associations and the prognosis model for sepsis will help physicians in ICUs to treat the disease.

Interesting findings were reported, including that a series of independent and significant prognosis factors were associated with the outcome. The associations involved age, GCS, SOFA, sepsis shock, breath rate, ALB, TBIL, CHE, BUN, LAC, OI, HCO3^-^, and IMV. ULR and MLR analyses controlling for confounders showed 13 factors independently and significantly associated with the outcome. The GCS score was used to evaluate the patient’s nervous system function as a part of the SOFA score .The mechanisms of these dysfunctions may involve inappropriate immune–brain signaling, the deleterious production of NO, and an endocrine response that can increase the risk of death in septic shock [19-22]. TBIL was used to quantify the degree of organ dysfunction over the course of sepsis and used to assess and define liver dysfunction. Several studies proved that an abnormal TBIL leads to a poor outcome in sepsis and septic shock[23, 24].The pathophysiology of sepsis-induced liver dysfunction was partly explained, including hypoxic and cholestasis aspects [25].Some studies showed CHE to be related to the outcome [26].The mechanism is because the cholinergic system plays an important role in the immune response to sepsis [27].Acute kidney injure (AKI) was significantly associated with the outcome [28]. Generally, septic AKI is profoundly different from ischemic AKI, both in the experimental setting and in the clinic [28]. LAC is typically used to evaluate disease severity, treatment response, and prognosis. The traditional view was that the rise in serum lactate concentration in sepsis is, in part, due to an impaired lactate clearance [29].

An association study was conducted in1,395 sepsis patients to explore candidate factors associated with a 28-day mortality of sepsis, indicating that the mean arterial pressure(MAP) is associated with mortality in critically ill sepsis patients[30].A multicenter, retrospective, observational study carried out in a Czech ICU reported that central venous pressure, MAP, and LAC were independently and significantly associated with the in-hospital mortality of sepsis[31].Traditionally, these factors, including age and the increased burden of chronic health conditions, were important risk factors for severe sepsis[1, 32]. Compared with the general population, there was a high prevalence in individuals with COPD, cancer, chronic renal, liver disease, or DM. In addition, malnutrition, residence in long-term care facilities, and abnormalities in the immune response to infection were also critical risk factors[32].Our findings were consistent with the results of previous studies. Genetic risk factors significantly influenced the susceptibility to and outcomes of sepsis [33]. Moreover, multiple genes may interact with pathogens to influence the response and outcome of sepsis, such as the genes of tumor necrosis factor (TNF), Toll-like receptor (TLR)-1, and TLR-4[33, 34].Unfortunately, our study could not gain the genetic information to explore the associations.

Another interesting finding was that high-performance mathematical prognosis models were successfully developed. ULR and MLR analyses were used to develop best-fit MLR models consisting of independently and significantly associated factors. We have not only selected statistically, independently, and significantly associated factors, but also considered these factors in relation to clinical experience and emergency medicine knowledge for the development of a best-fit prognosis model. In model 2, the prognosis models had a high prognosis value for the mortality of sepsis (AUC = 0.885, Table 6).The HL statistic and accuracy of model 2 were 4.977 and 0.814, respectively, indicating the model were successfully developed and had a high predictive performance. Furthermore, we developed a prognosis model based on SOFA in model 3 with an AUC of 0.736, which was consistent with the results of another six studies [35]. Interestingly, the top AUC was 0.756 in the six models, indicating our models include variables that represent the prognosis model for the outcome. Moreover, the prognosis performance of model 2 was significantly higher than that of model 3, showing that model 2 based on GCS combined with separate SOFA parameters had a significantly greater predictive performance for models based on SOFA score. This is partly because the single SOFA score combines these variables with a variable, leading to a decreased predictive power to detect the outcome. In addition, the continuous variable was categorized as a categorical variable for model development, which is highly important to increase the power to detect the mortality of sepsis.

Furthermore, we developed low- and high-risk groups for predicting the outcome based on the two prognosis models. In model 2, sepsis patients in the low-risk group had only a 0.049 mortality of sepsis, while the proportion of sepsis patients in the low-risk group in the total sample was 0.455. The mortality of sepsis among patients in the high-risk group was 0.903 (0.127 for the total sample). The proportion of low-risk and high-risk groups was 0.582 in the total sample. The low-risk and high-risk groups could greatly help physicians in emergency ICUs to treat and predict the disease outcome, which could benefit the security and quality of clinical practice, particularly in hospitals of mainland China[35, 36]. If sepsis patients have high risk scores, physicians could have good evidence to discuss the status of the patients with their partners or relatives. Unfortunately, there was little literature to report on prognosis or risk models for predicting the mortality of sepsis in China. To the best of our knowledge, the prognosis model was one of the best performance models for outcomes in China. In the near future, we will program a model using a web or computer platform to aid physicians in decision making concerning the prognosis of sepsis in emergency ICUs.

There were some limitations in this study. First, in this work, data analyses differed from previous studies, partly due to handling and managing heterogeneous and multiple-source electronic medical information. The experience and ability of our group in the series studies could reduce the negative influence. Second, selection bias could not be eliminated in the present study based on the real-world study design. Fortunately, sepsis patients were collected from a multicenter, which can reduce bias.

## Conclusion

The study showed evidence of independent and significant factors associated with the mortality of sepsis, including age, GCS, SOFA, septic shock, breath rate, TBIL, CHE, BUN, LAC, OI, HCO_3_^-^, IMV, and ALB. High-performance prognosis models for the mortality of sepsis were developed.

## Declaration statement

Consent for publication

All authors read and approved the final manuscript.

## Ethics approval and consent to participate

The study was approved by Ethics Committee of the first hospital affiliated to South China University, Hunan, China and performed in accordance with the Declaration of Helsinki.

## Availability of data and material

The datasets generated and/or analyzed during the current study are not publicly available due to private information but are available from the corresponding author on reasonable request. Dataset are from the study whose authors may be contacted at Center of Bioinformatics and Biostatistics, Institutes of Integrative Medicine, Fudan University.

## Author’s contributions

Y.L and Q.K drafted the manuscript. J.L, T.J participated in the design of the study and performed the statistical analysis. Z.T conceived of the study, and participated in its design and coordination and helped to draft the manuscript. All authors read and approved the final manuscript.

## Acknowledgments

We thank the grant from Institutes of Integrative Medicine of Fudan University to support the study.

## Authors’ Information

Y.L was Department of emergence medicine, the first hospital affiliated to South China University, Hunan, China; J.L was Department of emergence medicine, the hospital affiliated to Jining medical University, Shandong, China; T.J was Department of emergence medicine, Huaihua people’s hospital, Hunan, China; Q.K and Z.T was Department of Integrative Medicine, Huashan Hospital, and Institutes of Integrative Medicine, Fudan University, Shanghai, China.

